# Usage-inspired Interactive Digital Pathology System for Pathology Instruction in Indonesia

**DOI:** 10.1101/2024.10.19.24314313

**Authors:** Ery Kus Dwianingsih, Dewa Nyoman Murti Adyaksa, Petr Walczysko, Auliana Hayu Kusumastuti, Jean-Marie Burel, Jason R. Swedlow

## Abstract

**Background:** COVID-19 shifted Indonesian education to remote learning. The Gadjah Mada University (UGM) Faculty of Medicine Public Health and Nursing (UGM FMPHN) struggled with studying tumor images remotely due to resource shortages. In parallel, the University of Dundee’s (UoD’s) OME team created OMERO for image management, inspiring UGM to use OMERO to develop GamaPath for web-based image viewing. This report details its implementation, effectiveness, and potential expansion to practical sessions and workshops in Indonesia.

**Methods:** Teaching slides were scanned in Indonesia and imported into OMERO at UoD. UGM students, residents, and clinicians used GamaPath for anatomical pathology training for several sessions between 2022 and 2024. GamaPath was also used for national continuing education sessions for pathologists held through 2023. Experiences and evaluations were collected via online surveys and results were assessed by modified MARuL scores.

**Results:** The UoD-UGM collaboration produced an application satisfying the initial need for remote teaching during COVID lockdown. However, after returning to in-person teaching, access to interactive training materials online was considered to be an essential part of effective instruction by students and faculty. The GamaPath application provided flexible access to interactive materials that enhanced the educational experience. Of 256 survey respondents, mean modified MARuL score for GamaPath web app was 40.92 (SD = 10.73) with a median of 41 (IQR = 34– 50). Among the 107 pathologists who used GamaPath for national continuing education, the majority gave it the highest possible rating for functionality, ease of use, and overall experience.

**Conclusions:** UGM’s FMPHN collaborated with UoD to create a digital pathology platform based on OMERO, improving image analysis and feedback for practical sessions and workshops using GamaPath.

## Introduction

Like many locations worldwide, the COVID-19 pandemic transformed education in Indonesia, causing an abrupt and difficult shift to remote learning for safety. Schools quickly adapted to distance learning to maintain education services.^1,2^ Before March 2020, instruction of anatomical pathology students at Gadjah Mada University used traditional microscopes, with students directly viewing individual slides. The pandemic halted hands-on training, and initially, instruction was provided using single-field images in PowerPoint for Zoom lectures. This had the obvious limitations of incomplete coverage and compromised resolution. The result was a degraded experience for both faculty and students.

The sudden change highlighted a key aspect of pathology training in Indonesia and most likely in other low- and middle-income countries (LMICs). Modern clinical practice relies on robust pathology services, requiring access to labs and trained personnel. Expert staff are fundamental, offering diagnosis and therapeutic advice, particularly in LMICs where local disease knowledge is vital, so developing appropriate training programs to develop and maintain expertise in pathology is crucial. However, LMICs face resource limitations in delivering updated pathology training. Standard textbooks often lack locally relevant disease information. Digital pathology, involving digitally scanned Whole Slide Images (WSIs), offers a solution but presents challenges. High costs, limited availability of slide scanners, and the need for high-performance computing (HPC) systems complicate implementation.^3^

These deficits are more than inconveniences. The United Sustainable Development Goals (UN SDGs) 3 (Good Health and Well-Being) and 4 (Quality Education) prioritize education and healthcare access for robust societies. At UGM, over 50% of anatomical pathology students are women. Access to quality education is essential for developing highly trained female clinicians (UN SDG 5, Gender Equality). The pandemic disrupted education, particularly affecting the training of expert clinical staff and women severely.^4^

To address this challenge, we established an intercontinental collaboration between FMPHN UGM (Yogyakarta, Indonesia) and the University of Dundee (UoD, Dundee, UK) that provided digital pathology instruction for UGM students and that has now become an Indonesian national resource. At FMPHN UGM, there are no slide scanners nor advanced HPC facilities, but an internationally recognized medical school provides clinical training. At UoD, the Open Microscopy Environment (OME) team has developed OMERO, an open-source image data management system that provides web-based WSI viewing.^5^ OMERO offers secure image data sharing over a standard network connection. The collaboration provided a long-distance pathology learning method for UGM students and faculty using digital pathology. This initiative was called the GamaPath project.

In this report, we describe the work to date on GamaPath as a tool for instruction in anatomy and pathology, evaluate its use and utility by UGM students, and extend its use to regional and national case review workshops for pathologists across Indonesia.

## Methods

### Technical Solution

#### Slide Scanning

We sent a set of teaching slides to Jakarta for whole slide scanning using an Aperio CS2 from Leica Biosystems Imaging, Inc., Leica Microsystems (SEA) Pte Ltd (Singapore, Jakarta, Indonesia) (Figure 1). The slide scanner operates on the Microsoft Windows 10 operating system, captures slide images from the desktop, and creates high-quality digital slides. It has a five-slide capacity and 20X and 40X magnification capabilities, making it a highly reliable solution for medium-volume sites. The Aperio CS2’s intuitive interface and easy-to-use design consistently deliver rapid creation of high-resolution digital slides, with a >98% first-time scan success rate. After the scanning of the slides was completed, all the WSIs were transferred back to UGM using external hard drives and then uploaded to the OMERO platform via OMERO.importer.^5^ The imported images were organized in OMERO based on their label image and the name or requirements of the virtual lab session. Login credentials, lasting from three days to one month, are rotated after each virtual session.

**Figure 1.**
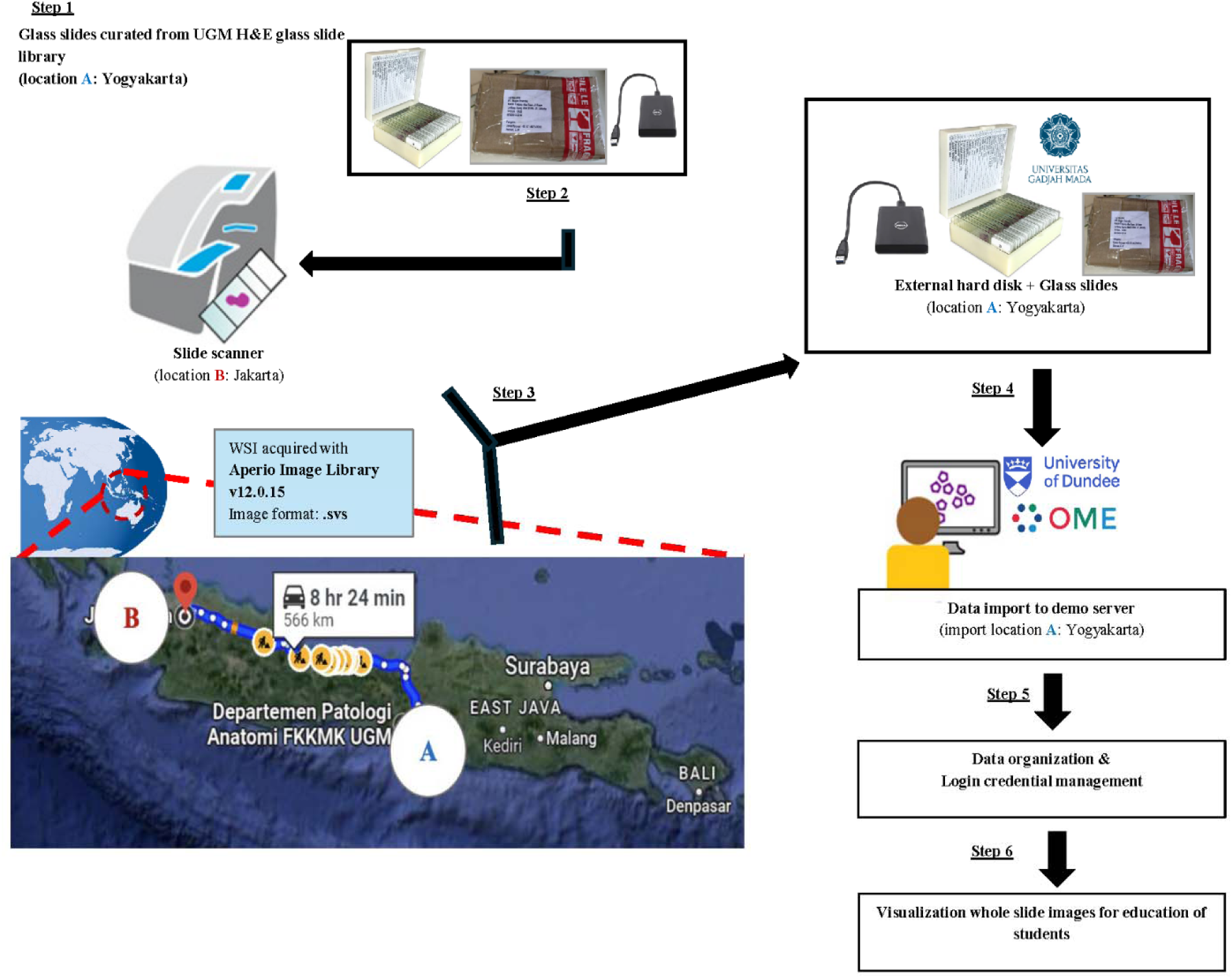
GamaPath-OMERO Workflow System The curated slides library was collected from the Department of Anatomical Pathology at FMPN UGM (location A, in Yogyakarta). The teaching glass slides are stained with Hematoxylin-eosin (H&E) and various immunohistochemical stains. The slides were carefully packed to prevent breakage and driven to Jakarta along with an external hard drive for the whole slide scanning process (location B). They were then scanned using the Aperio Digital Pathology Slide Scanner CS2. The scanned results were stored on a hard drive and then returned alongside the slides to location A. The WSI files in SVS file format were imported into an OMERO running at UoD. Initially, we used the OMERO demo server, but a dedicated resource was made available in September 2023. The imported data were organised and annotated to supplement the undergraduate students’ educational journey and provide valuable resources for pathology residents to enhance their training and knowledge.

#### Web-based Access

GamaPath uses OMERO, a client-server application for storing and managing imaging data. The Bio-Formats library^6^ reads WSIs in SVS format, supporting a multi-resolution format. GamaPath’s OMERO web client offers scalable WSI viewing and web-based access for students and educators. Benefits include practical slide storage, easy maintenance, and improved user experience, enabling remote classroom work. WSI technology allows users to choose magnification without specific technical skills, unlike traditional microscopes.^7,8^

#### Friendly User Interface

OMERO’s web interface offers an interactive, visually engaging user experience. It allows users to explore microscopic digital images, with options to zoom in and out and select specific features. This platform facilitates efficient study and research with customization options. Students, particularly novices, can intuitively adapt to digital images for a seamless experience. Clinical medicine has also validated Digital Slides from Virtual Microscopes for histopathology diagnosis, showing no significant difference from traditional slides.^9,10^

#### Open Source

The high costs of WSI scanners and Virtual Microscope software hinder the adoption of digital microscopy in medical education at UGM. The key for GamaPath was system reconfigurability to meet UGM faculty and student needs and integrate curriculum. We incorporated open-source software for UGM customization, like adding their logo, organizing data by modules, and creating an access structure for resource management. Using OMERO supports transparency, collaboration, and the inclusion of UGM’s developments in future updates. This approach reduces costs and supports integration with other educational tools.

#### Flexibility

During the COVID-19 pandemic, educators were forced to transition to an online distance learning pedagogy.^11,12^ GamaPath is designed to allow flexibility in deployment and usage. This adaptability, hopefully, supports diverse teaching methods and accommodates varying levels of technological infrastructure within educational settings in Indonesia.

#### Remote Hosting

Students have returned to in-person learning, but many pandemic innovations like remote learning are being integrated into everyday practice.^13^ Digitizing the workspace has improved learning. A survey found pathology students using Digital Slide in a remote clerkship reported increased interest and understanding.^14^ Installing an OMERO system needs IT resources and staff, which are not always available. Thus, the OME team offers a free server, the OMERO Demo server, at UoD for evaluation. During the GamaPath initiative’s initial phase, UGM used UoD’s OMERO Demo server, loading 235 WSIs (519 GB). Despite the limitations of the Demo server, UGM trained students and clinicians. In late 2023, UoD provided dedicated IT resources for a customized OMERO platform for UGM. This helped UGM manage and expand digital pathology resources efficiently, offering accessibility and scalability.

#### Cost-Effective

By utilizing open-source software and adopting remote hosting strategies, GamaPath offers a cost-effective and sustainable solution for digital pathology education. This is critical for resource-constrained institutions seeking to enhance educational offerings without significant financial investments in proprietary technologies or infrastructure.

### Evaluation of the Web-Apps value using Modified MARuL Score

In this study, undergraduate students and residents filled out the modified MARuL (Mobile App Rubric for Learning) questionnaire during practical sessions. The MARuL scoring system is a quick and user-friendly method that teachers can use to evaluate the value of an app for just-in-time learning.^15^ This rubric comprises items in four categories: Part A (Teaching & Learning Measures), Part B (User-Centered Measures), Part C (Professional Measures), and Part D (Usability Measures). These criteria represent the value of just-in-time learning as assessed by students. The MARuL category score is calculated by adding the rating for each item on a 5- point Likert-type scale (0 = does not fulfill the item requirements, 1 = poorly fulfills the requirements, 2 = somewhat fulfills the requirements, 3 = mostly fulfills the requirements, and 4 = fully meets the requirements) within each category to reach a total score for that category (teaching and learning = 36, user-centered = 28, professional measures = 12, and usability = 28). Adding up the scores of the categories gives the user an overall score of 104. Apps are then categorized by their scoring range (< 50 = not at all valuable, 51-69 = potentially valuable, and > 69 = probably valuable).^15^

Our study omitted Part C (Professional Measures) of the MARuL score because the respondents were students. The three categories used were teaching and learning (n = 3), user-centered (n = 7), and usability (n = 4). In this study, the modified MARuL category score was calculated by adding the rating for each item on a 5-point Likert-type scale within each category to reach a total score for that category (teaching and learning = 12, user-centered = 28, and usability = 16. Summing the categories gives the user an overall score of 56 (Supplementary Table S1). Apps are then categorized by their scoring range (< 25 = not at all valuable, 25-35 = potentially valuable, and > 35 = probably valuable).

In addition to conducting a practical session for students, a workshop was organized for pathologists. The User Feedback Score was utilized to evaluate the functionality, ease of use, and overall experience of the GamaPath web application for anatomical pathology training. This questionnaire consists of 5-point Likert-type scale (1 = very dissatisfied, 2 = dissatisfied, 3 = neutral, 4 = satisfied, and 5 = very satisfied) (Supplementary Table S2).

## Results

The timeline of our project is shown in Figure 2. FMPHN UGM has driven the GamaPath project, beginning with digital slide scanning and then defining and implementing WSI sharing in UGM’s anatomical pathology department using OMERO. The project aligns with the school’s vision of becoming a world-class, innovative, and superior faculty of medicine by optimizing the use of data and information technology to serve the nation’s and humanity’s interests. Therefore, the project has received four years of consecutive limited funding, allowing the FMPHN UGM team to evaluate aspects of the implementation to date, including the practical session for undergraduates and residents and a professional workshop for pathologists. FMPHN UGM’s use of OMERO started with a test account running on the OMERO Demo Server provided by the UoD-based OME team. FMPHN UGM handled all configuration, delivery, and evaluation of GamaPath using the Dundee-based OMERO Demo server from Q2 2022 to Q4 2023, ensuring the system was tuned to its needs.

**Figure 2.**
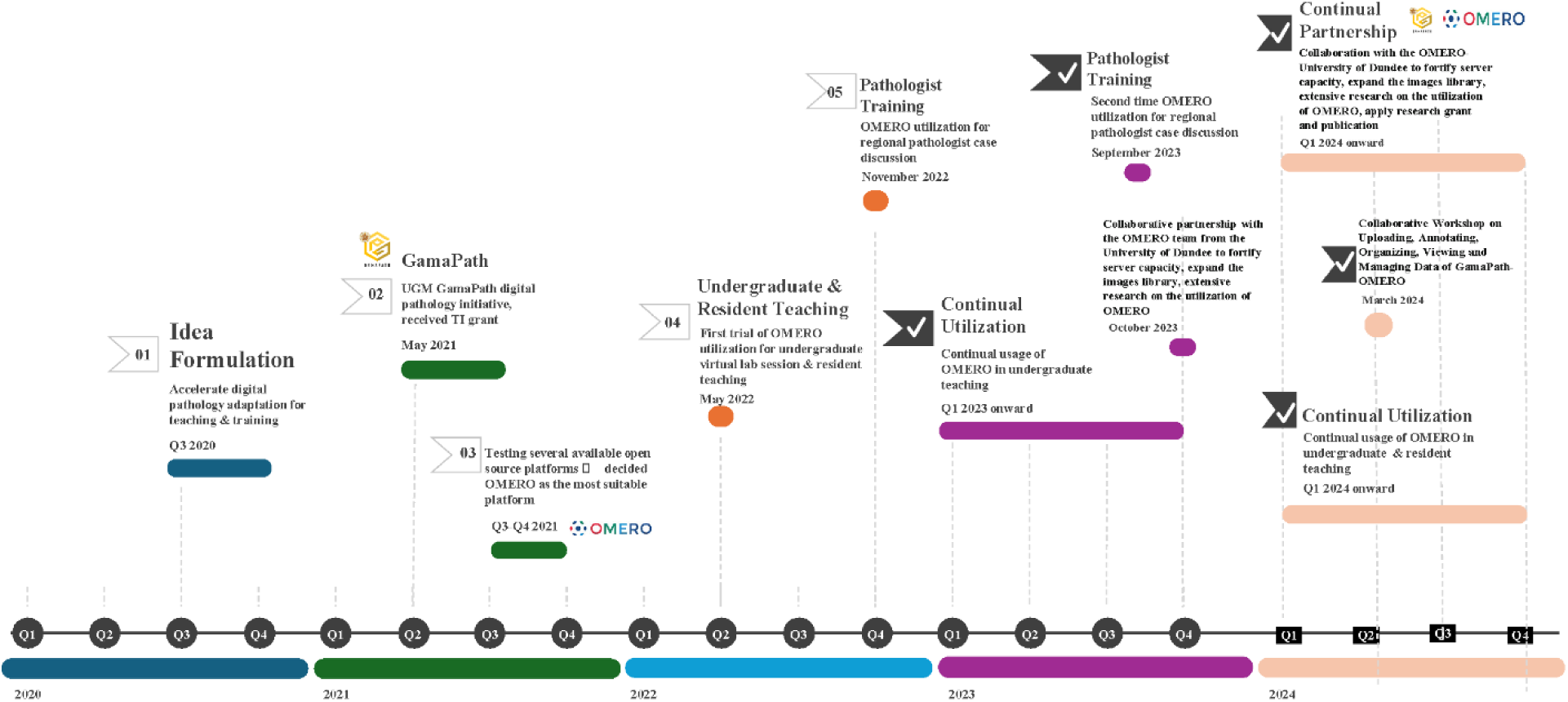
GamaPath-OMERO Project Development Timeline. The idea for the GamaPath-OMERO project was formulated in the third quarter of 2020 to accelerate the adaptation of digital pathology for teaching and training purposes. In the following year, GamaPath digital pathology was initiated at UGM and received University support in the form of a small grant in May 2021. After testing several available open-source platforms from the third to the fourth quarter of 2021, OMERO was chosen as the most suitable platform for GamaPath. The first trial of OMERO utilization for undergraduate virtual lab sessions and resident teaching began in May 2022. Following the success of the first trial, GamaPath- OMERO was then used for pathologist training in November 2022, marking the first utilization for regional pathologist case discussions. Additionally, GamaPath-OMERO saw its second utilization in September 2023 for regional pathologist case discussions. In October 2023, collaborative partnership with the OMERO team from UoD was established to improve the server capacity, expand the image library, conduct extensive research on the utilization of OMERO, and apply for research grants and publications. In March 2024, a collaborative workshop on uploading, annotating, organizing, viewing, and managing data of GamaPath- OMERO was held at Universitas Gadjah Mada (UGM). Then, the utilization of OMERO in undergraduate and resident teaching will continue from 2024 onward.

The GamaPath user interface is shown in Figure 3. Sets of WSI datasets are organized and annotated using the OMERO.web interface (Fig. 3A). Different sets of WSIs, selected for each of the FMPHN UGM pathology courses are displayed in different folders in the left-hand panel of the browser interface. Users open a fully interactive viewing module (Fig. 3B) by double-clicking on the WSI names or thumbnails in the center panel. The web browser UI application connects to an OMERO server, which stores and serves all data and configurations^5^.

**Figure 3.**
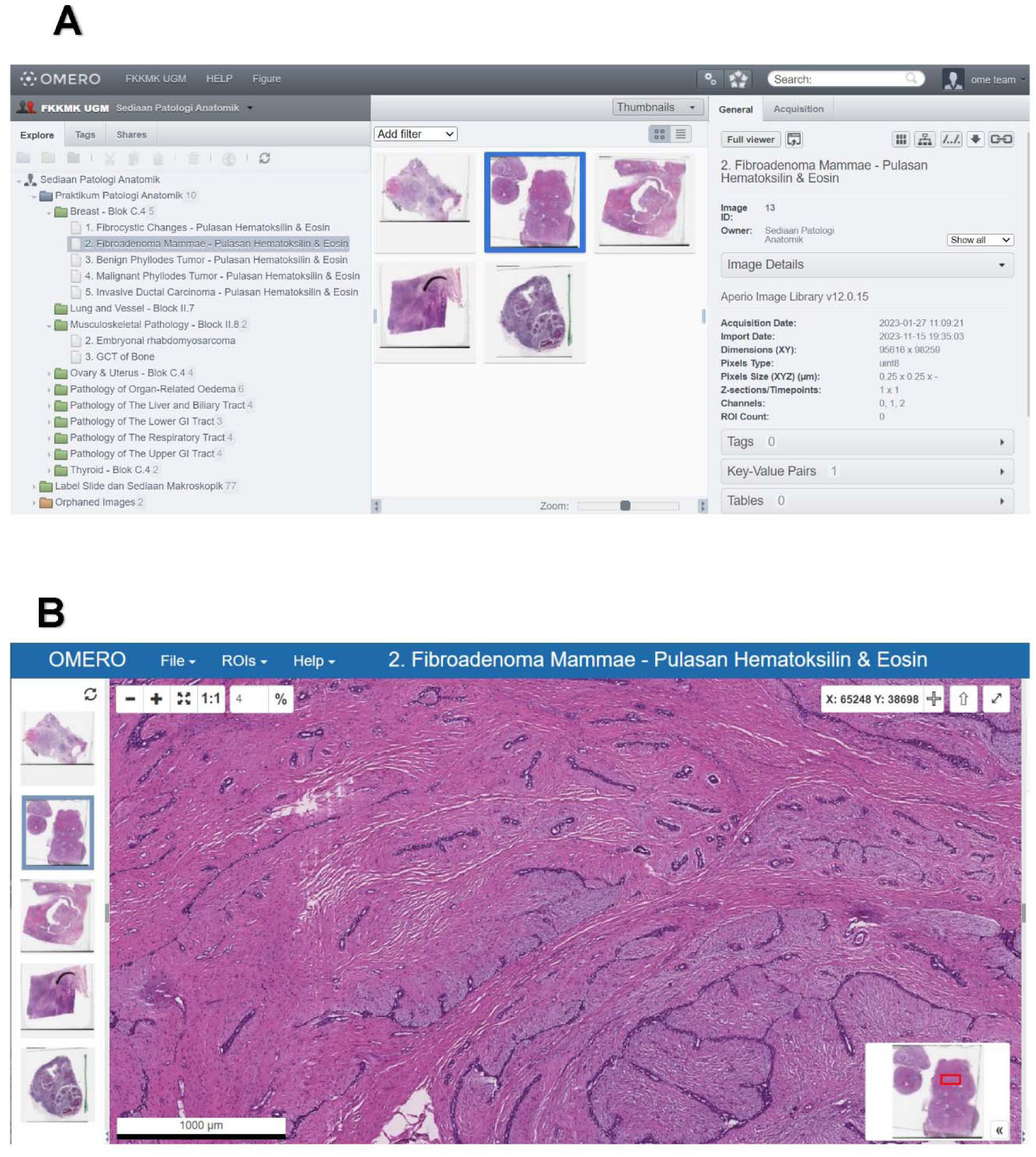
GamaPath User Workflow and Interface A. The screenshot shows the typical layout of a virtual microscopy lab into separate foilders (projects and datasets in OMERO.web (see Allan et al (2012). Each department (anatomical pathology, histology, etc.) or organ system pathology (musculoskeletal, neuropathology, etc.) is assigned to different members of a specific group for students or pathologists. Dataset content i tailored based on the learning objectives required in each student/course module. The screenshot shows the typical layout of an OMERO.iviewer after double-clicking on an image thumbnail in the central panel of OMERO.web (A). The left panel of the viewer gives the user access to all WSIs within a dataset for easy comparison of different slides and cases.

The GamaPath platform runs on a virtual machine with 100GB RAM, 16 cores, and 1.5TB storage, with data backed up at UoD and a local WSI copy at UGM. To date, 911 GB of data (60% more than the Demo server) has been imported. The data trains UGM undergraduates (72 slides), residents (78 slides), and clinicians across Indonesia (218 slides, 517 GB). Clinicians accessed 26 slides in the 2022 symposium, 51 more in 2023, and will access 141 new slides in Q4 2024 (Table 1). The new OMERO server has been active since January 2024, with over 3400 logins in the first six months, indicating GamaPath’s growing importance for UGM (Figure 4).

**Figure 4.**
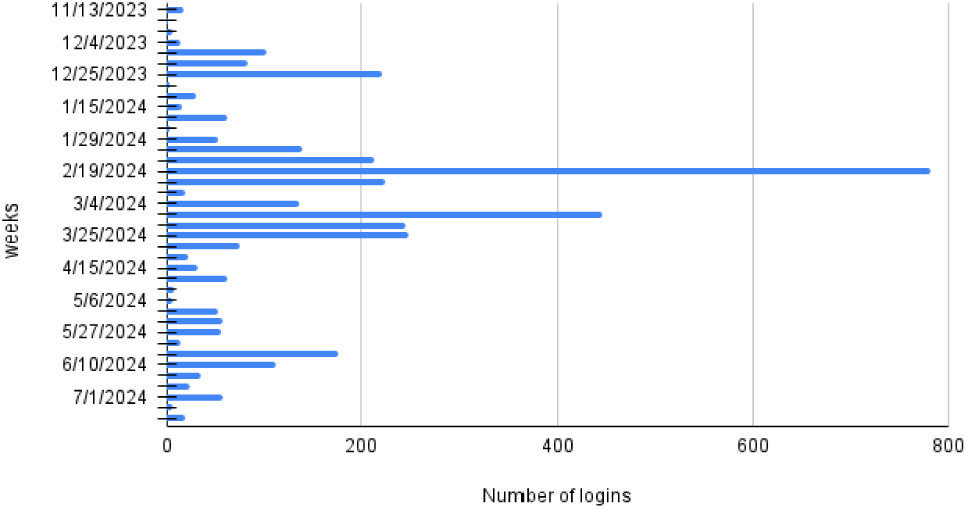
Number of Logins to GamaPath-OMERO Plot shows the number of logins per week. The platform is used regularly by residents and clinicians. The spikes correspond to either teaching blocks or access prior to assessments.

**Table 1.** Distribution of The Slides Used to Train Undergraduate and Residents.

| Course | Total number of slides | Access by Undergraduate | Access by Residents |
| --- | --- | --- | --- |
| Histology | 71 (82 GB) | First and Second year: 41 slides | 30 slides extra |
| Anatomical Pathology | 79 (380 GB) | Second and third year: 31 slides | 38 slides extra |

From a total of 71 slides (82 GB) for the histology course, 41 slides were by the first and second undergraduate students, and 30 extra slides were accessed by residents. From a total of 79 slides (380 GB) for the anatomical pathology course, 31 slides were accessed by the second and third undergraduates, and 38 extra slides were accessed by residents.
Abbreviations: GB – Gigabyte.

Once the first version of GamaPath was validated, a formal collaboration began between the UGM and UoD teams. This resulted in the provision of an OMERO system at UoD dedicated to FMPHN UGM’s use, with scalable storage and computing resources (see Methods). This approach was selected because while UGM had a small server available on its campus, the number of WSIs needed to make GamaPath useful for students and faculty required additional storage capacity. However, with the success of the GamaPath project, we aim to migrate the system to UGM-based systems once they are available.

During daily practices, anatomical pathology residents at the FMPHN UGM encounter cases at Sardjito General Hospital, a UGM-affiliated teaching hospital. Residents initially interpret the cases, which consultant pathologists review, revise, and verify. Some cases are routine, and others may be challenging, requiring confirmation through additional examinations like immunohistochemical analysis or PCR, for instance, spindle cell sarcoma or pleomorphic sarcoma. Rare cases, such as adamantinoma of the bone, may also be encountered. By collecting these cases, scanning them, and incorporating them into GamaPath, the morphological images of those cases can be preserved and studied by generations of residents regardless of whether they encounter them in daily practice.

### Introduction and Evaluation of GamaPath and OMERO

During the practical session, the instructor demonstrated techniques for using and visualizing microscopic digital images in GamaPath. This was followed by a lecture on various tumor entities, focusing on digital images, like ductal carcinoma of the breast. Students then independently explored and analyzed the images to deepen their understanding. At the end of the session, all students completed a modified MARuL questionnaire to provide feedback on their experiences with GamaPath.

Among 256 practical session respondents, 82 were male and 174 female. In this study, the modified MARuL score for teaching and learning measures was 8·8008 (SD 2·56434), with a median score of 9 (IQR = 7–11). The modified MARuL score result for user-centered measures was 20·3711 (SD 5·54686) with a median score of 21 (IQR = 16·25–25·0). The modified MARuL score result for usability measures was 11·7500 (SD 3·20049) with a median score of 11·75 (IQR = 9·25–14·0) (Table 2).

**Table 2.**
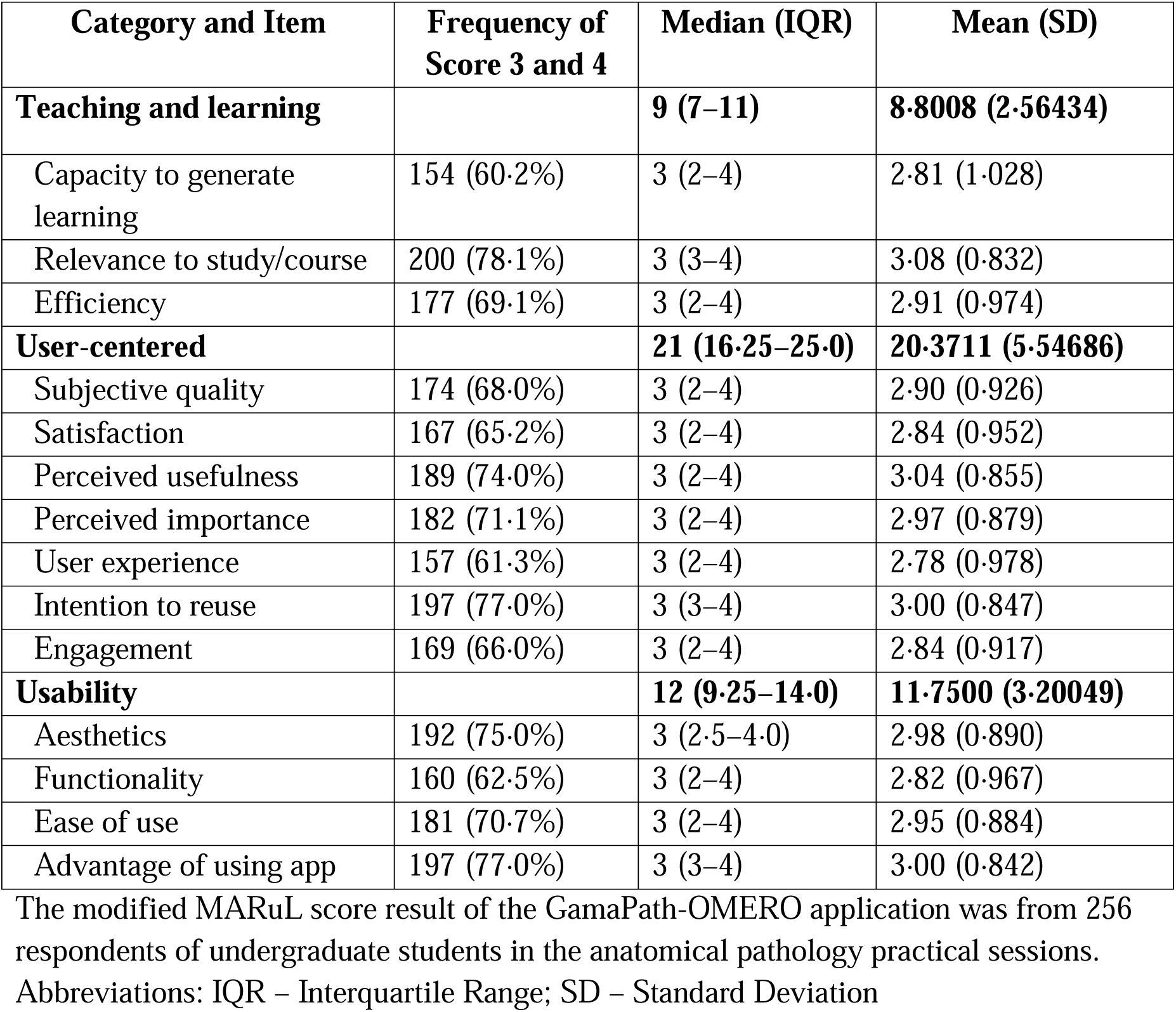
The Modified MARuL Score Result of GamaPath-OMERO Application.

| Category and Item | Frequency of Score 3 and 4 | Median (IQR) | Mean (SD) |
| --- | --- | --- | --- |
| **Teaching and learning** |  | **9 (7–11)** | **8.8008 (2.56434)** |
| Capacity to generate learning | 154 (60.2%) | 3 (2–4) | 2.81 (1.028) |
| Relevance to study/course | 200 (78.1%) | 3 (3–4) | 3.08 (0.832) |
| Efficiency | 177 (69.1%) | 3 (2–4) | 2.91 (0.974) |
| **User-centered** |  | **21 (16.25–25.0)** | **20.3711 (5.54686)** |
| Subjective quality | 174 (68.0%) | 3 (2–4) | 2.90 (0.926) |
| Satisfaction | 167 (65.2%) | 3 (2–4) | 2.84 (0.952) |
| Perceived usefulness | 189 (74.0%) | 3 (2–4) | 3.04 (0.855) |
| Perceived importance | 182 (71.1%) | 3 (2–4) | 2.97 (0.879) |
| User experience | 157 (61.3%) | 3 (2–4) | 2.78 (0.978) |
| Intention to reuse | 197 (77.0%) | 3 (3–4) | 3.00 (0.847) |
| Engagement | 169 (66.0%) | 3 (2–4) | 2.84 (0.917) |
| **Usability** |  | **12 (9.25–14.0)** | **11.7500 (3.20049)** |
| Aesthetics | 192 (75.0%) | 3 (2.5–4.0) | 2.98 (0.890) |
| Functionality | 160 (62.5%) | 3 (2–4) | 2.82 (0.967) |
| Ease of use | 181 (70.7%) | 3 (2–4) | 2.95 (0.884) |
| Advantage of using app | 197 (77.0%) | 3 (3–4) | 3.00 (0.842) |

The modified MARuL score result of the GamaPath-OMERO application was from 256 respondents of undergraduate students in the anatomical pathology practical sessions.
Abbreviations: IQR – Interquartile Range; SD – Standard Deviation

Figure 5A–C show the distribution of scores in the different evaluation categories. Students rated GamaPath highly and overall found that it enhanced their learning. The distributions of all measured features confirmed the value of the application. The Kolmogorov-Smirnov test (*p* = 0·000, *p* < 0·05) showed non-normal data (Supplementary Figure S1, Supplementary Figure S2).

**Figure 5.**
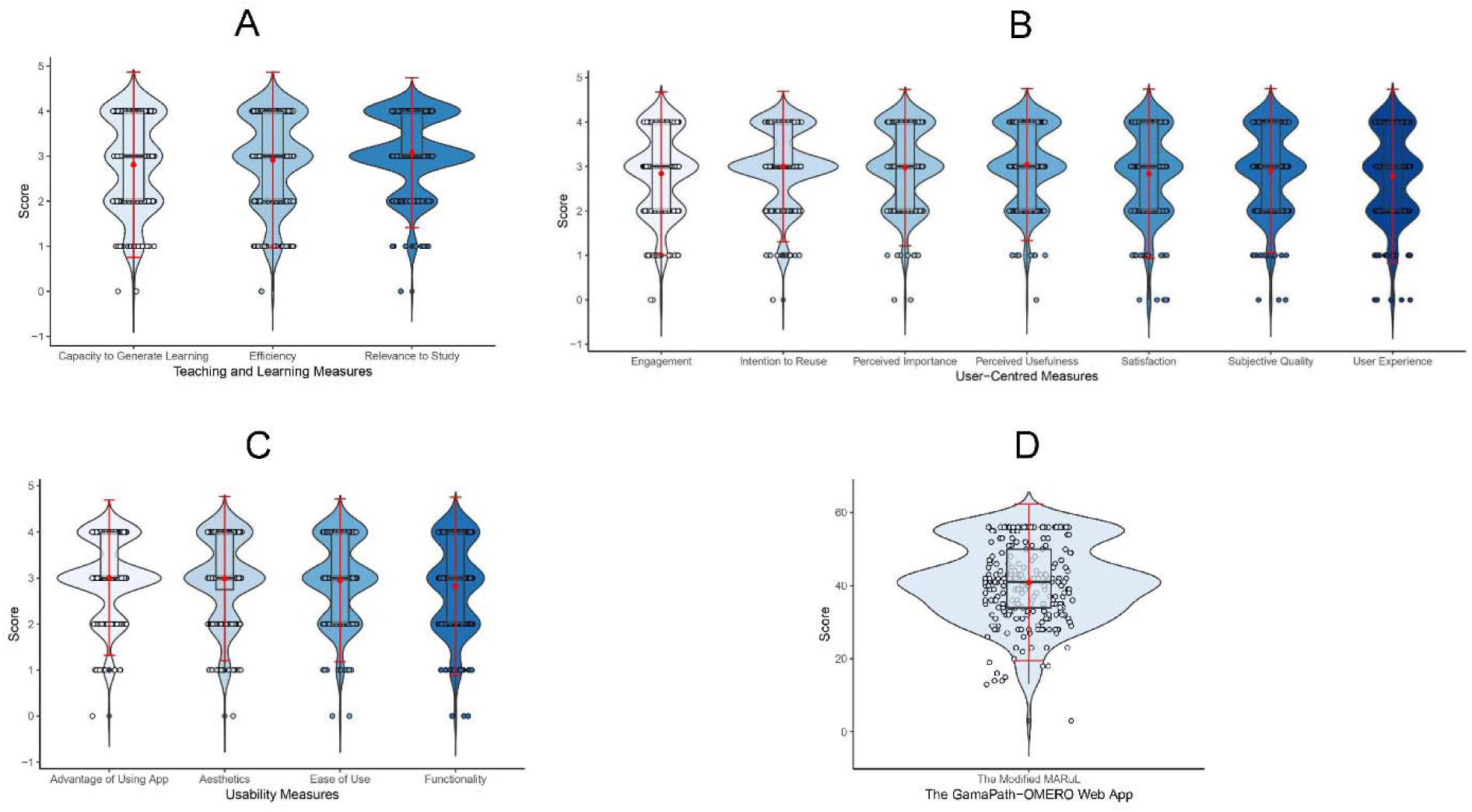
Summary of Student Survey Responses. Violin plots for (A) Teaching and learning measures, (B) User-centered measures, (C) Usability measures, and (D) Modified MARuL Score Result. Violin plots are hybrids of boxplots and density plots used here to visualize the distribution of modified MARuL score results. The thick band represents the 50th percentile, the thinner bands represent the 25th and 75th percentiles. The red dot represents the mean (40·92), the red line represents the standard deviation (10·729), and the black thick band represents the median (41).

The mean of the modified MARuL score was 40·92 (SD = 10·729), with a median of 41 (IQR = 34–50) (Figure 5D, Supplementary Table S3), indicating that the application should be considered as “probably valuable”. Scores ranged from 3 to 56 (Supplementary Table S4, Supplementary Figure S3). We conclude that the GamaPath application is providing substantial benefit to students but that further improvements and development should be undertaken to make the application as useful as possible.

### Using GamaPath for National Case Review

Besides the practical session for undergraduates and residents, a workshop for professional pathologists in Indonesia was also held to assess the system’s suitability for them. Participants joined the workshop via Zoom and viewed the slides presented through GamaPath, all directly connecting to the OMERO server based at UoD. This was especially vital considering Indonesia’s geography, enabling participants from Java, Sumatra, Sulawesi, and Borneo islands to participate and benefit from the training. During the workshop, the instructor delivered a detailed lecture using PowerPoint. The digital slides of tumor cases from the GamaPath library were also discussed with the participants. Subsequently, pathologists had the opportunity to individually examine and analyze the digital images of each tumor type, allowing them to gain practical experience and enhance their understanding of the subject. At the workshop, pathologists were requested to complete a user feedback questionnaire to gather valuable insights regarding their experiences and feedback using GamaPath.

Out of a total of 120 pathologists who participated in the workshop, of which more than 50% were female, 107 pathologists gave their feedback as follows: 63 respondents (58·9%) rated the functionality aspect with a score of 5 out of 5, 52 respondents (48·6%) rated the ease of use aspect with a score of 5 out of 5, and 49 respondents (45·8%) provided a score of 5 out of 5 for the overall experience of the GamaPath web application (Table 3). The Kolmogorov-Smirnov test (*p* = 0·000, *p* < 0·05) showed non-normal data (Supplementary Figure S4). The mean of user feedback score result was 4·56 (SD 0·552) with a median score of 5 (IQR = 4–5) for the functionality aspect, 4·36 (SD 0·706) with a median score of 4 (IQR = 4–5) for the ease of use, and 4·36 (SD 0·664) with a median score of 4 (IQR = 4–5) for the overall experience (Table 3, Figure 6A). The mean of the user feedback score was 13·29 (SD = 1·754) with a median of 13 (IQR = 12–15) (Figure 6B; Supplementary Table S5). Scores ranged from 7 to 15 (Supplementary Table S6, Supplementary Figure S5).

**Figure 6.**
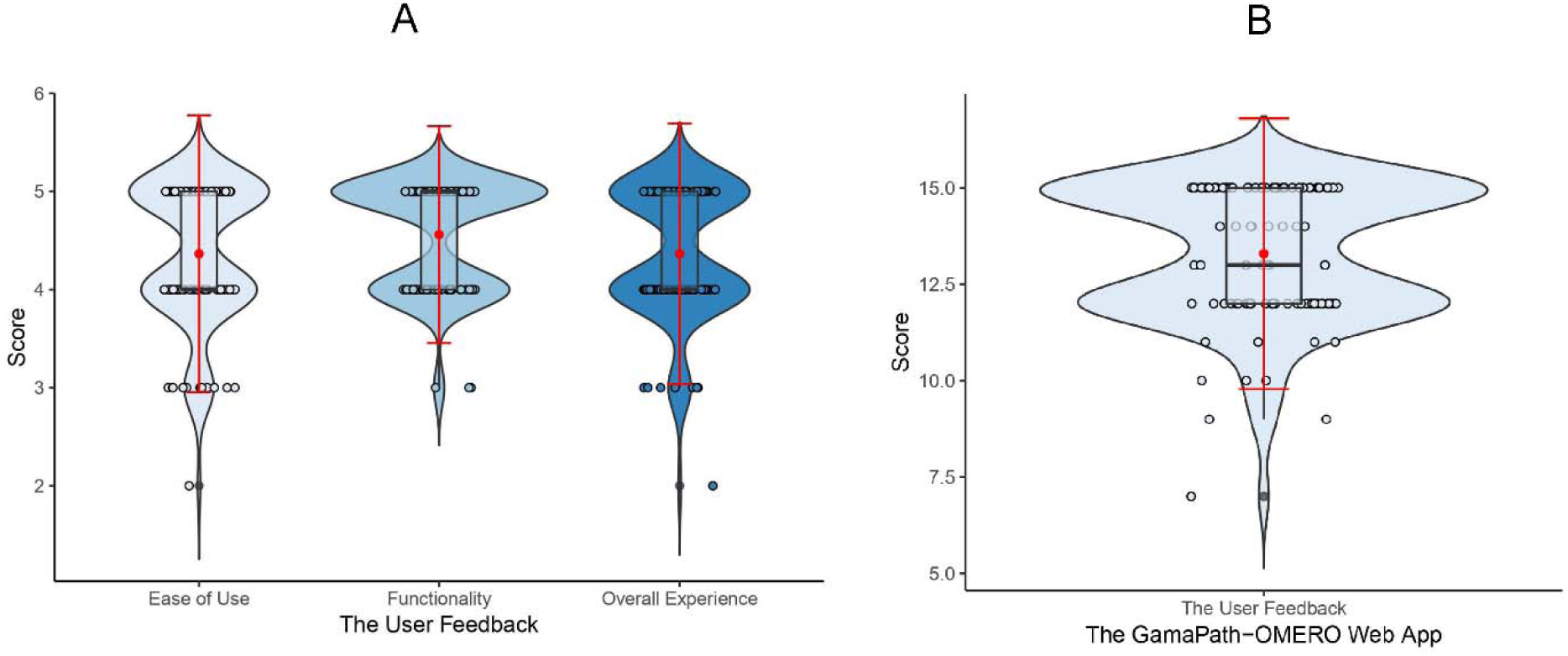
Summary of Pathologist Survey Responses. Violin plots for (A) Ease of Use, Functionality, and Overall Experience, and (B) The Pathologist’s User Feedback Score Result. Violin plots are hybrids of boxplots and density plots used here to visualize the distribution of user feedback score results. The thick band represent the 50th percentile, the thinner bands represent the 25th and 75th percentiles. The red dot represents the mean (13·29), the red line represents the standard deviation (1·754), and the black thick band represents the median (13).

**Table 3.** The User Feedback Score Result of GamaPath-OMERO Implementation for Anatomical Pathology Training.

| Category | Frequency of Score 5 | Median (IQR) | Mean (SD) |
| --- | --- | --- | --- |
| Functionality | 63 (58.9%) | 5 (4–5) | 4.56 (0.552) |
| Ease of Use | 52 (48.6%) | 4 (4–5) | 4.36 (0.706) |
| Overall Experience | 49 (45.8%) | 4 (4–5) | 4.36 (0.664) |

Out of a total of 120 pathologists who participated in the workshop, of which more than 50% were female, 107 pathologists gave their feedback.
Abbreviations: IQR – Interquartile Range; SD – Standard Deviation

## Discussion

We have designed, constructed, and successfully run a widely accessible digital training solution for sharing WSIs and training undergraduate students, residents, and pathologists in Indonesia called GamaPath. The solution leverages open-source software and strategic collaboration between UoD and UGM, which have partnered to exchange expertise, know-how, and technology resources to benefit the respective institutions’ communities. GamaPath wa originally built to address the teaching conditions caused by the COVID-19 pandemic. However, the strong acceptance and adoption of GamaPath by all stakeholders has driven the continuation and growth of the project. At the time of writing, over 360 WSI datasets have been stored in GamaPath and used for instruction and review by several hundred students, residents, and pathologists at UGM and across Indonesia.

GamaPath was originally designed to address a critical need for remote instruction but has rapidly grown far beyond its initial scope and aims. This was possible because of the commitments of UGM and UoD to identify paths for collaboration as well as the open-source nature of the OMERO software that is used as its foundation. By using an open solution, UGM’s project team could decide how to configure the system to best deliver on their goals. Discussions between the UGM and UoD technical teams on the configuration of OMERO proceeded with both teams speaking from a position of knowledge, which allowed rapid and creative solutions to emerge.

Originally focused on UGM’s needs, GamaPath has expanded its impact, aligning with several UN Sustainable Development Goals: SDG 3 (health workforce), SDG 4 (affordable quality education and technical skills for youth), SDG 5 (women’s participation and empowerment through technology), and SDG 17 (international cooperation and knowledge sharing). These alignments emerged from the project’s collaborative and open-source approach. This is consistent with the foundations of the principles of open-source distribution^16^ and provides an example of how open-source technology and transnational institutional collaboration contribute towards the delivery of the UN SDGs.

In March 2024, the ongoing collaboration between UGM and UoD led to a collaborative workshop held at FMPHN UGM, dedicated to exploring the full potential of the GamaPath platform. The workshop was attended by members of the Anatomical Pathology Department staff and residents, as well as representatives from various other departments, including Histology, Parasitology, and Microbiology. Notably, a representative from the Faculty of Dentistry also took part in the workshop. The goal of the workshop was to delve into the intricacies of uploading, annotating, organizing, viewing, and managing data, with the ultimate aim of maximizing the platform’s capabilities in the context of FMPHN UGM.

This initiative empowered FMPHN at UGM to lead digital pathology education without requiring extensive resources. Using the OMERO Demo server, FMPHN UGM conducted workshops for students, residents, and pathologists, yielding impressive outcomes. A strategic partnership with UoD enabled the provision of a larger server, expanding the digital repository and long-term data storage. FMPHN UGM now excels in digitizing anatomical pathology resources, enhancing its education system, and attracting students locally and regionally, solidifying UGM’s reputation. For UoD, the collaboration increases engagement with international partners, develops equitable partnerships, and promotes joint research and capacity building in health and education systems using GamaPath.

To measure mobile learning potential in clinical skills, users should evaluate an app’s value in supporting student learning. MARuL and user feedback scores help users assess an app’s effectiveness. These scores provide a structured framework for evaluating an app, ensuring it meets educational needs and contributes to clinical skills development.^15,20^ Assessing the modified MARuL score during practical sessions and the user feedback score in the pathologist workshop for the GamaPath Web platform showed its significant value. Further evaluation and optimization of GamaPath’s usability are essential for user experience and effectiveness, benefiting educational and professional applications.

Using dedicated IT resources, we expanded GamaPath’s whole slide images (WSI), enabling FMPHN UGM to store a wide range of cases. These cases support laboratory sessions and workshops and include rare ones, ensuring long-term access and continuity in medical education. This provides invaluable resources for students and practitioners, exposing them to diverse pathological conditions. WSI in digital pathology is used for distance teaching and AI-based research, such as protein expression quantification and mutation detection.^17–19^ These advances will further enhance UGM’s digital pathology capabilities.

## Contributors

EKD and DNMA conceptualized and designed the study and were supported in executing the study by PW, JMB, and JRS. DNMA, PW, and JMB led data collection and import. DNMA and AHK were responsible for the data cleaning, analysis, and interpretation. AHK, PW, and JMB prepared tables and figures. EKD wrote the first draft of the manuscript. All authors contributed substantially to the critical review, editing, and revision of the manuscript. All authors had full access to all data in the study, approved the final version of the manuscript, and had final responsibility for the decision to submit the manuscript for publication.

## Data Sharing

The data in this study is available from the Supplementary Info.

## Data Availability

All data produced in the present work are contained in the manuscript

## Acknowledgments

The development of digital pathology at UGM was supported by the Technology and Information Grant (Hibah Teknologi Informasi) provided by the UGM Faculty of Medicine, Public Health, and Nursing to EKD. The UoD-based OME team was supported by an award from The Wellcome (Ref.: 221361/Z/20/Z). Travel funds for the UoD team were funded by the Scottish Education Exchange Programme Test and Learn Project.

