## Supplementary Information for "Usage-inspired Interactive Digital Pathology System for Pathology Instruction in Indonesia"

**Appendix A: Supplementary Tables**

Supplementary Table S1. Modified MARuL Questionnaire

Supplementary Table S2. User Feedback Questionnaire for The GamaPath–OMERO Web Application

Supplementary Table S3. The Modified MARuL Score Result

Supplementary Table S4. The Frequency of Modified MARuL Score

Supplementary Table S5. The User Feedback Score Result
Supplementary Table S6. The Frequency of User Feedback Score

**Appendix B: Supplementary Figures**

Supplementary Figure S1. Normality Test Result of The Modified MARuL Score

Supplementary Figure S2. Normality Test Result Post Exclusion of Outlier Data

Supplementary Figure S3. The Frequency of the Modified MARuL Score Result

Supplementary Figure S4. Normality Test Result of The User Feedback Score

Supplementary Figure S5. The Frequency of the User Feedback Score Result

**Appendix A: Supplementary Tables**

**Supplementary Table S1. Modified MARuL Questionnaire**

| **MODIFIED MARUL QUESTIONNAIRE** | | | | | | |
| --- | --- | --- | --- | --- | --- | --- |
| **Teaching and learning measures** | | | | | | |
| **No** | **Questions** | **0** | **1** | **2** | **3** | **4** |
| 1 | **Capacity to generate learning:** Does GamaPath support the achievement of your learning goals as outlined in the study guide? |  |  |  |  |  |
| 2 | **Relevance to study/course:** Is the information on this app relevant to your study needs? Does it align with the course material? |  |  |  |  |  |
| 3 | **Efficiency:** Would GamaPath increase the effectiveness of your learning? Would it make it easier and quicker for you to learn? |  |  |  |  |  |
| **User-centered measures** | | | | | | |
| 1 | **Subjective quality:** Does the GamaPath seem like a good app? Would you recommend the app to the other medical students? |  |  |  |  |  |
| 2 | **Satisfaction:** Am I happy with the app's performance? Would GamaPath help motivate you and help you achieve your study goals in this module/block? |  |  |  |  |  |
| 3 | **Perceived usefulness:** Does the app seem like it would be useful and help you with your learning? |  |  |  |  |  |
| 4 | **Perceived importance:** Does the app appear to be an important part of your learning? Does it look like a resource that would improve your study routine? |  |  |  |  |  |
| 5 | **User experience:** Does the app perform the way I expect it to? |  |  |  |  |  |
| 6 | **Intention to reuse:** After trying the app (GamaPath Web-OMERO), do you think you would use it again? |  |  |  |  |  |
| 7 | **Engagement:** Is the app interesting, fun, and stimulating? |  |  |  |  |  |
| **Usability measures** | | | | | | |
| 1 | **Aesthetics:** Does the app's design, interface, information presentation, graphics, and layout help or hinder its use? |  |  |  |  |  |
| 2 | **Functionality:** How well does the app work in terms of accuracy, ease of performing tasks, speed, and performance? |  |  |  |  |  |
| 3 | **Ease of use:** How easy is it to learn how to use the app? How clear are the icons/menus and instructions? |  |  |  |  |  |
| 4 | **Advantage of using app over web-based or conventional equivalent:** Is there an advantage to using the app over a resource on the internet or a textbook? For example, interactivity and portability? |  |  |  |  |  |

**Supplementary Table S2. User Feedback Questionnaire for The GamaPath–OMERO Web Application**

| **User Feedback Questionnaire for The GamaPath–OMERO Web Application** | | | | | |
| --- | --- | --- | --- | --- | --- |
| **Question** | **Answer** | | | | |
| How useful is this digital pathology application? | Very unuseful | Unuseful | Neutral | Useful | Very Useful |
|  | 1 | 2 | 3 | 4 | 5 |
| How easy this application can be used? | Very difficult | Difficult | Neutral | Easy | Very easy |
|  | 1 | 2 | 3 | 4 | 5 |
| How is your overall experience about this application? | Very dissatisfied | Dissatisfied | Neutral | Satisfied | Very Satisfied |
|  | 1 | 2 | 3 | 4 | 5 |

**Supplementary Table S3. The Modified MARuL Score Result**

| **Modified MARuL Score Result** | | | |
| --- | --- | --- | --- |
|  | | **Statistic** | **Standard Error** |
| Mean | | 40.92 | 0.671 |
| 95% Confidence Interval for Mean | Lower Bound | 39.60 |  |
|  | Upper Bound | 42.24 |  |
| 5% Trimmed Mean | | 41.43 |  |
| Median | | 41.00 |  |
| Variance | | 115.115 |  |
| Standard Deviation | | 10.729 |  |
| Minimum | | 3 |  |
| Maximum | | 56 |  |
| Range | | 53 |  |
| Interquartile Range | | 16 |  |
| 25th Percentile | | 34 |  |
| 75th Percentile | | 50 |  |
| Skewness | | -0.346 | 0.152 |
| Kurtosis | | -0.099 | 0.303 |

The modified MARuL score result of the GamaPath-OMERO application from 256 respondents of undergraduate students in anatomical pathology practical session showed scores ranging for 3–56 with a mean score of 40.92 (SD = 10.729), a median score of 41, and an interquartile range of 16 from 34–50.

**Supplementary Table S4. The Frequency of Modified MARuL Score**

| **Modified MARuL Score** | | | | |
| --- | --- | --- | --- | --- |
| Score | Frequency | Percent | Valid Percent | Cumulative Percent |
| 3 | 1 | 0.4 | 0.4 | 0.4 |
| 13 | 1 | 0.4 | 0.4 | 0.8 |
| 14 | 2 | 0.8 | 0.8 | 1.6 |
| 15 | 1 | 0.4 | 0.4 | 2.0 |
| 16 | 1 | 0.4 | 0.4 | 2.3 |
| 18 | 2 | 0.8 | 0.8 | 3.1 |
| 19 | 1 | 0.4 | 0.4 | 3.5 |
| 20 | 1 | 0.4 | 0.4 | 3.9 |
| 22 | 1 | 0.4 | 0.4 | 4.3 |
| 23 | 4 | 1.6 | 1.6 | 5.9 |
| 26 | 2 | 0.8 | 0.8 | 6.6 |
| 27 | 3 | 1.2 | 1.2 | 7.8 |
| 28 | 13 | 5.1 | 5.1 | 12.9 |
| 29 | 5 | 2.0 | 2.0 | 14.8 |
| 30 | 2 | 0.8 | 0.8 | 15.6 |
| 31 | 6 | 2.3 | 2.3 | 18.0 |
| 32 | 10 | 3.9 | 3.9 | 21.9 |
| 33 | 7 | 2.7 | 2.7 | 24.6 |
| 34 | 8 | 3.1 | 3.1 | 27.7 |
| 35 | 9 | 3.5 | 3.5 | 31.3 |
| 36 | 9 | 3.5 | 3.5 | 34.8 |
| 37 | 5 | 2.0 | 2.0 | 36.7 |
| 38 | 6 | 2.3 | 2.3 | 39.1 |
| 39 | 10 | 3.9 | 3.9 | 43.0 |
| 40 | 7 | 2.7 | 2.7 | 45.7 |
| 41 | 14 | 5.5 | 5.5 | 51.2 |
| 42 | 24 | 9.4 | 9.4 | 60.5 |
| 43 | 10 | 3.9 | 3.9 | 64.5 |
| 44 | 5 | 2.0 | 2.0 | 66.4 |
| 45 | 5 | 2.0 | 2.0 | 68.4 |
| 46 | 5 | 2.0 | 2.0 | 70.3 |
| 47 | 5 | 2.0 | 2.0 | 72.3 |
| 48 | 3 | 1.2 | 1.2 | 73.4 |
| 49 | 3 | 1.2 | 1.2 | 74.6 |
| 50 | 3 | 1.2 | 1.2 | 75.8 |
| 51 | 4 | 1.6 | 1.6 | 77.3 |
| 52 | 3 | 1.2 | 1.2 | 78.5 |
| 53 | 5 | 2.0 | 2.0 | 80.5 |
| 54 | 2 | 0.8 | 0.8 | 81.3 |
| 55 | 5 | 2.0 | 2.0 | 83.2 |
| 56 | 43 | 16.8 | 16.8 | 100.0 |
| Total | 256 | 100.0 | 100.0 |  |

The modified MARuL score result of the GamaPath-OMERO application from 256 respondents showed that the lowest score was 3 and the highest score was 56.

**Supplementary Table S5. The User Feedback Score Result**

| **Modified MARuL Score Result** | | | |
| --- | --- | --- | --- |
|  | | **Statistic** | **Standard Error** |
| Mean | | 13.29 | 0.170 |
| 95% Confidence Interval for Mean | Lower Bound | 12.95 |  |
|  | Upper Bound | 13.63 |  |
| 5% Trimmed Mean | | 13.43 |  |
| Median | | 13.00 |  |
| Variance | | 3.076 |  |
| Standard Deviation | | 1.754 |  |
| Minimum | | 3 |  |
| Maximum | | 15 |  |
| Range | | 8 |  |
| Interquartile Range | | 3 |  |
| 25th Percentile | | 12 |  |
| 75th Percentile | | 15 |  |
| Skewness | | -0.732 | 0.234 |
| Kurtosis | | 0.280 | 0.463 |

The User Feedback score result of the GamaPath-OMERO application from 107 respondents of pathologists in the national case review workshop showed scores ranging for 3–15 with a mean score of 13.29 (SD = 1.754), a median score of 13, and an interquartile range of 3 from 12–15.

**Supplementary Table S6. The Frequency of User Feedback Score**

| **User Feedback Score** | | | | |
| --- | --- | --- | --- | --- |
| Score | Frequency | Percent | Valid Percent | Cumulative Percent |
| 7 | 1 | 0.9 | 0.9 | 0.9 |
| 9 | 2 | 1.9 | 1.9 | 2.8 |
| 10 | 3 | 2.8 | 2.8 | 5.6 |
| 11 | 4 | 3.7 | 3.7 | 9.3 |
| 12 | 36 | 33.6 | 33.6 | 43.0 |
| 13 | 8 | 7.5 | 7.5 | 50.5 |
| 14 | 8 | 7.5 | 7.5 | 57.9 |
| 15 | 45 | 42.1 | 42.1 | 100.0 |
| Total | 107 | 100.0 | 100.0 |  |

The User Feedback score result of the GamaPath-OMERO application from 107 respondents showed that the lowest score was 7 and the highest score was 15.

**Appendix B: Supplementary Figures**

**Supplementary Figure S1. Normality Test Result of The Modified MARuL Score**

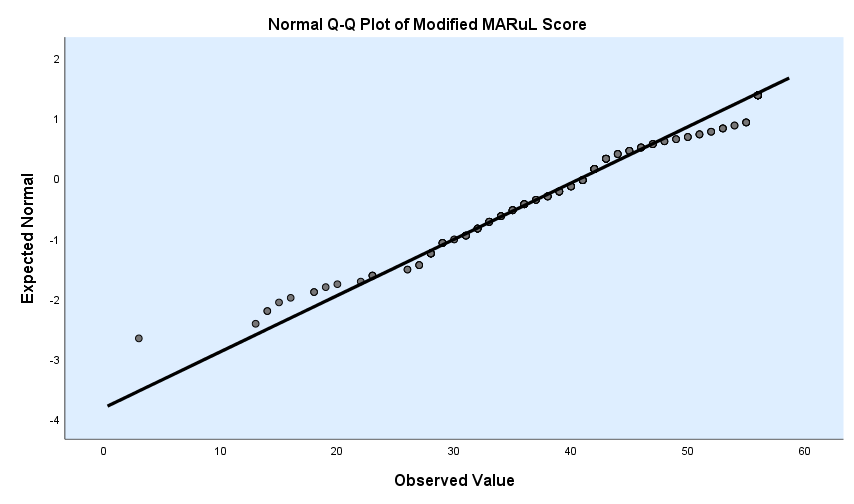

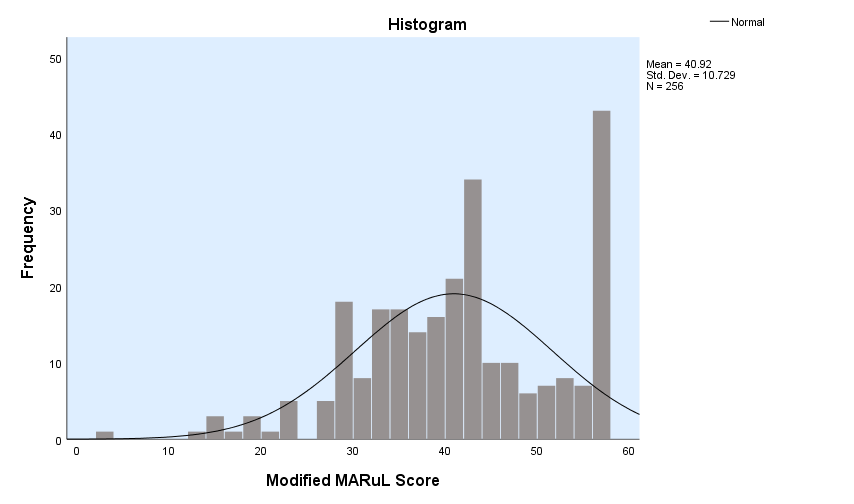

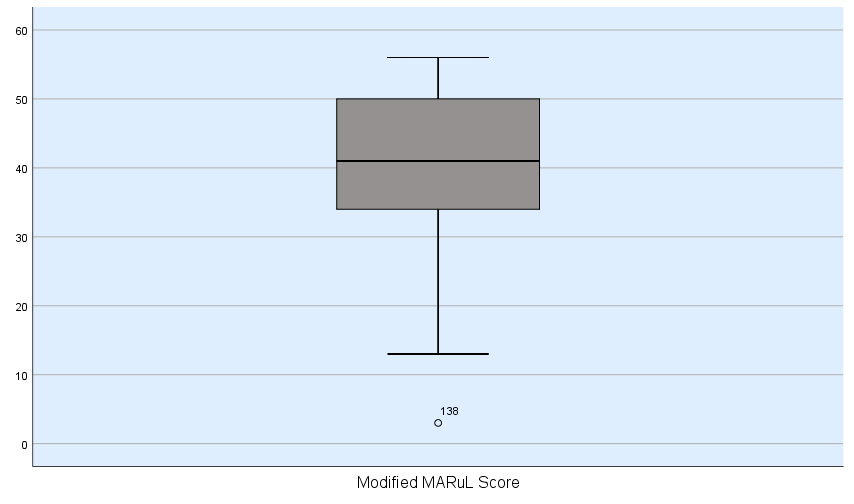

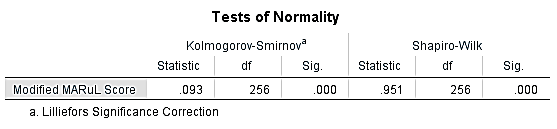

The Q-Q plot, histogram, box plot, and the Kolmogorov-Smirnov test showed *p* value 0.000 (*p* <0.05). The modified MARuL score result was not normally distributed.

**Supplementary Figure S2. Normality Test Result Post Exclusion of Outlier Data**

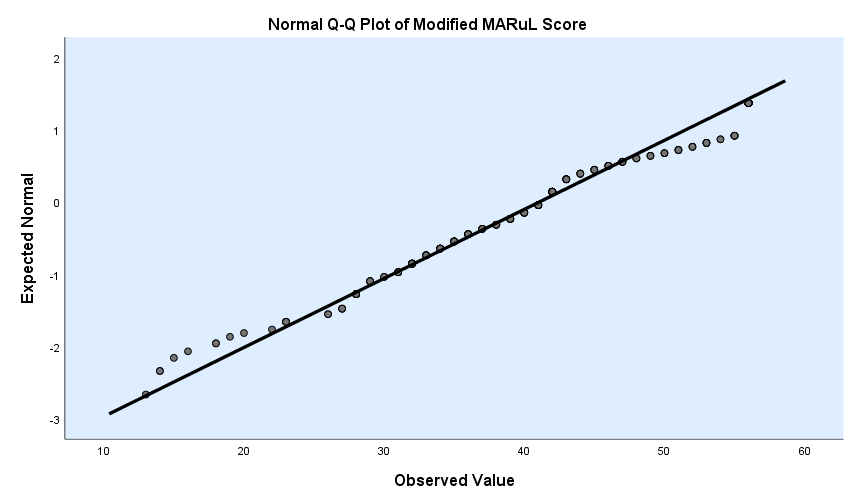

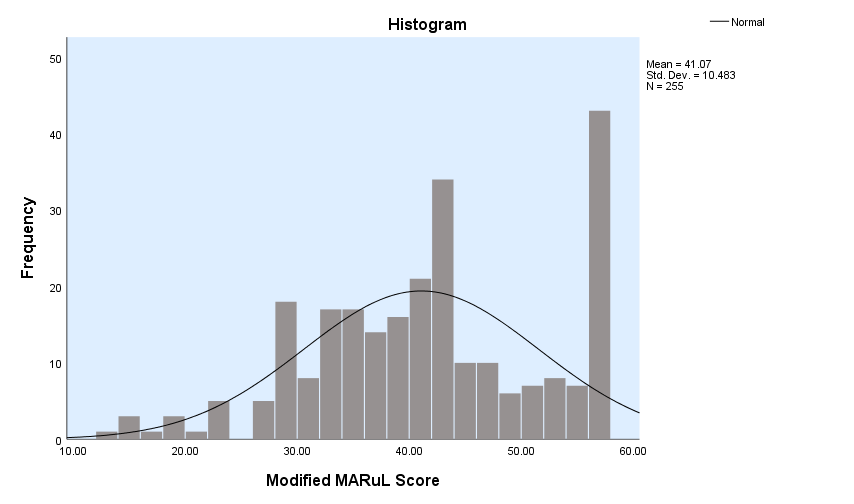

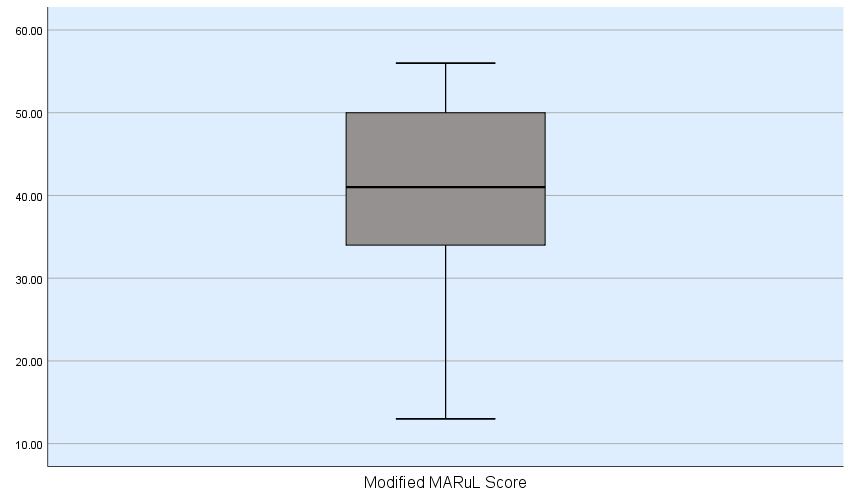

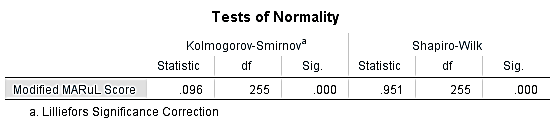

The Q-Q plot, histogram, box plot, and the Kolmogorov-Smirnov test post exclusion of outlier data showed *p* value 0.000 (*p* <0.05). The modified MARuL score result post exclusion of outlier data was not normally distributed.

**Supplementary Figure S3. The Frequency of the Modified MARuL Score Result**

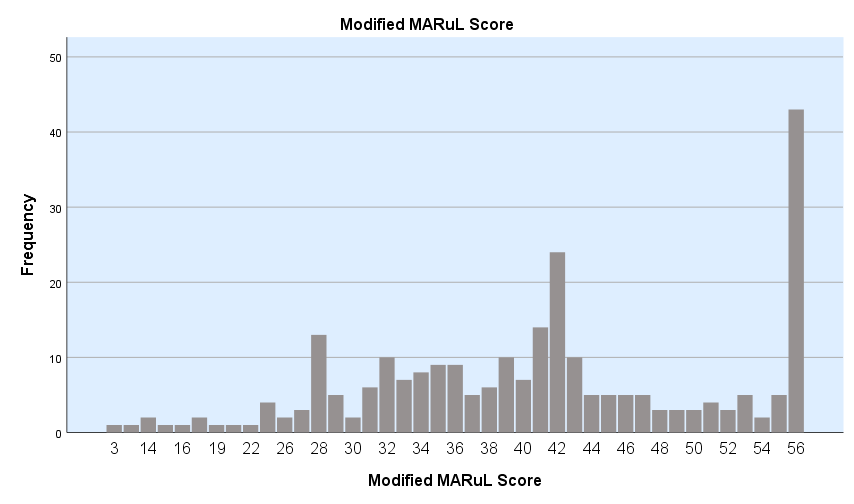

There were 43 respondents (16.8%) gave score 56 as the highest score and 1 respondent (0.4%) gave score 3 as the lowest score for the application.

**Supplementary Figure S4. Normality Test Result of The User Feedback Score**

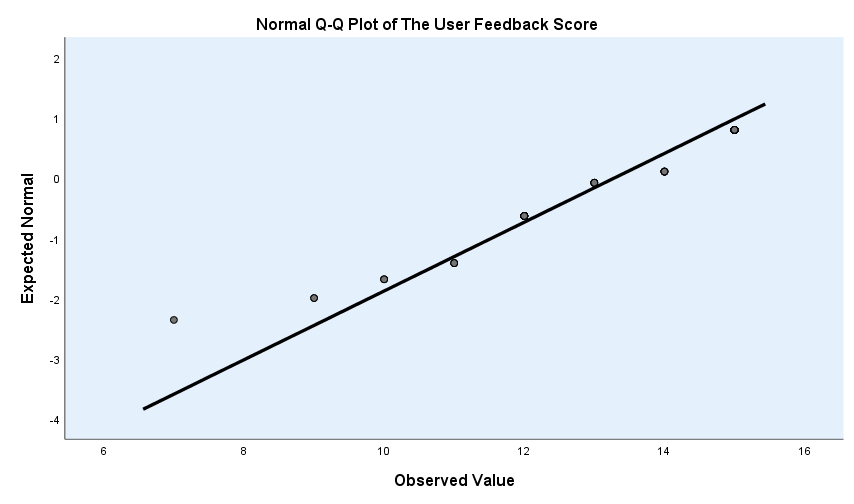

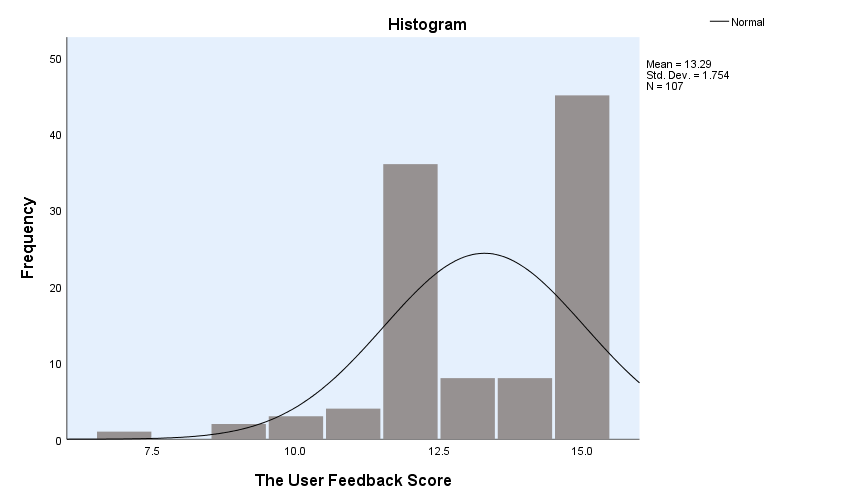

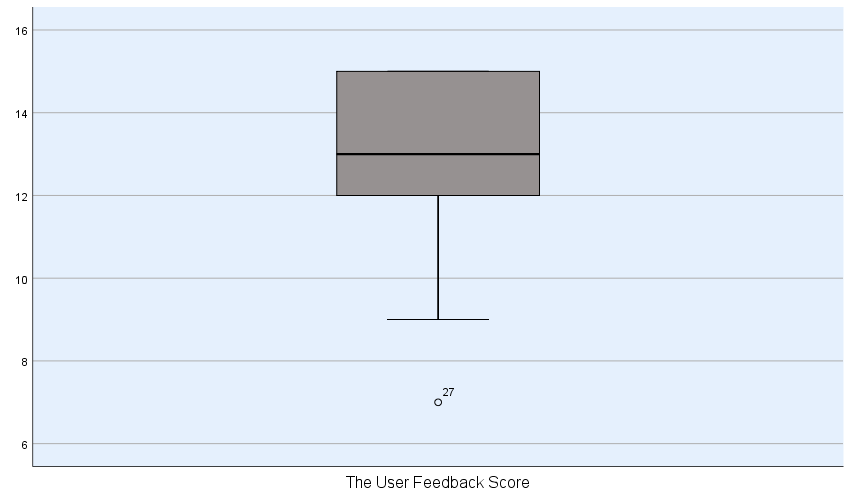

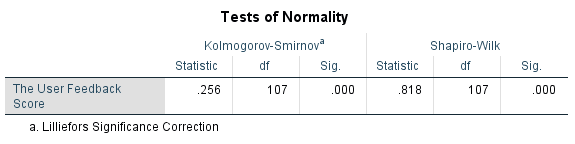

The Q-Q plot, histogram, box plot, and the user feedback score showed *p* value 0.000 (*p* <0.05). The user feedback score result was not normally distributed.

**Supplementary Figure S5. The Frequency of the User Feedback Score Result**

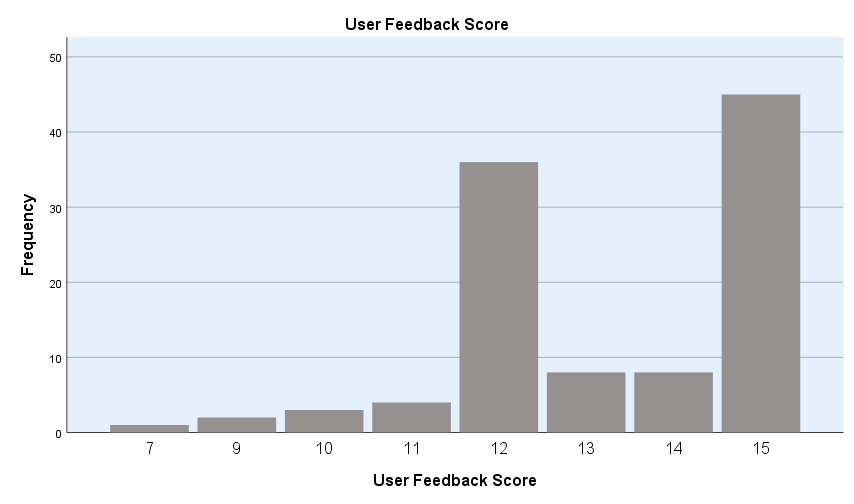

There were 45 respondents (42.1%) gave score 15 as the highest score and 1 respondent (0.9%) gave score 7 as the lowest score for the application.
